# Electroencephalographic insights into variant function and clinical outcomes in *SCN2A* encephalopathy

**DOI:** 10.1101/2023.10.24.23296360

**Authors:** Michelle Fogerson, Melina Tsitsiklis, Elise Brimble, Tobias Brünger, Alexander R. Arslan, Jayne Nerrie, Kim P. Laberinto, Alix M.B. Lacoste, Richard L. Martin, Dennis Lal, Jay Pathmanathan, M. Brandon Westover, Nasha Fitter, Jacob Donoghue

## Abstract

Neither phenotype nor genotype reliably predicts clinical disease severity or neurodevelopmental outcomes in *SCN2A* developmental and epileptic encephalopathy. In this study we examined the electroencephalographic (EEG) features of children with *SCN2A* variants to quantify the range of EEG abnormalities and link EEG biomarkers to developmental outcomes.

We retrospectively analyzed data from a cohort of 28 children with *SCN2A* variants and employed a genetics-based consensus framework to infer the functional characterization of each subject’s *SCN2A* variant. Eleven subjects were predicted to have a gain-of-function variant, and 17 subjects a loss-of-function variant. Overall, variant classifications matched subject phenotypes. 493 EEG recordings from the 28 subjects were analyzed (ages 1 day to 16 years). In addition to the *SCN2A* recordings, normative data from 1230 children without an epilepsy diagnosis or epileptiform features based on neurologists’ review was analyzed (1704 EEG recordings, ages 0 days to 16 years). We detected interictal epileptiform discharges (IEDs) in the *SCN2A* recordings using Beacon’s automated IED detection algorithm. We characterized background spectral features by computing relative power in four frequency bands (delta=1-4Hz, theta=4-8Hz, alpha=8-13Hz, beta=13-30Hz) in recordings from both the *SCN2A* and control cohorts. Additionally, we determined whether each *SCN2A* recording was associated with a gross motor developmental delay based on reported attainment of gross motor milestones. We then used mixed effects logistic regression models to estimate the effect of EEG biomarkers on developmental delay.

We characterized EEG abnormalities in the background spectral features of the *SCN2A* cohort compared to the controls and identified differences in EEG signatures between the subjects with gain- and loss-of-function variants. Additionally, we showed that background spectral features are correlated with motor developmental outcome when measured relative to age-matched neurotypical children. Furthermore, we showed that interictal epileptiform activity is correlated with delayed motor development in subjects with gain-of-function variants.

Taken together, these findings suggest that EEG biomarkers can be used to identify neurological abnormalities that correlate both with *SCN2A* variants and measures of development. We demonstrate the potential value of EEG as a disease biomarker, and we highlight the potential of such biomarkers to both guide future invasive genetic therapies and to be used as diagnostic tools.

## Introduction

Voltage-gated sodium ion channels, essential to the generation and propagation of bioelectrical signals, are encoded by a diverse but highly conserved family of genes. Each channel is formed from a single alpha subunit protein combined with one or more beta subunits.^1^ The relationship between neurological disease and variants in the genes encoding the components of sodium channels was first confirmed in 1998, with the identification of disease-causing variants in *SCN1B* causing generalized epilepsy with febrile seizures.^2^ However, disease-causing variants of the main alpha subunit have proven to be far more common and significant causes of human morbidity, and in 1999 a locus on chromosome 2 harboring the *SCN1A*, *SCN2A*, and *SCN3A* was identified.^3^ These genes express alpha subunits with both differing kinetics and patterns of expression.

By 2001, the general relationship between *SCN2A* variants and human disease was established. Initially, *SCN2A* variants were described in association with mild or “benign” febrile and nonfebrile seizures^4^ but soon after were linked to more severe neurological disease. Two broad phenotypes have emerged, the first involving variants resulting in early life seizures and often severe epileptic encephalopathies,^5–7^ and the second involving developmental delay with autism spectrum disorders.^8,9^ This dichotomy has been associated with the changes to the functional properties of the *SCN2A* alpha subunit at critical developmental stages. This subunit underpins the Na_v_1.2 channel, which is expressed in fetal life along the nodes of Ranvier and axon initial segments (AIS) of cortical glutamatergic pyramidal neurons (although it is subsequently retained only at the AIS).^8,10–12^ These findings suggest a role for Na_v_1.2 channels in both electrical signaling and neuronal network formation^8^. Gain-of-function (GoF) variants (increasing sodium conductance) tend to lead to epileptic disorders, while loss-of-function (LoF) variants (reducing sodium conductance) lead to autism and developmental delay with or without clinical seizures and epilepsy.^7,8,13^

Despite the relationship between phenotype and channel conductance, these properties fail to reliably predict clinical disease severity or neurodevelopmental outcomes, as do genotype and most early life exam features.^14^ The success of emerging gene therapies^15^ relies on timely and accurate characterization of developmental course and treatment response. Quantitative neurobiomarkers hold potential to predict clinical trajectories and guide early interventions such that the promise for disease modification supersedes the risks of invasive treatments.

Electroencephalography (EEG) is known to capture background and ictal abnormalities in *SCN2A* developmental and epileptic encephalopathy (DEE), even in early life.^13,16,17^ EEG is also used to diagnose and grade many of the underlying epileptic syndromes associated with *SCN2A* (such as Ohtahara syndrome, epilepsy of infancy with migrating focal seizures, West syndrome, Dravet syndrome, and Lennox-Gastaut syndrome). Quantitative EEG biomarkers are therefore a promising avenue for measuring the diagnostic efficacy of *SCN2A*-DEE treatments under development. Here, we characterize the variants of a large *SCN2A*-DEE cohort to identify where they fall on the GoF to LoF spectrum using a novel consensus framework that combines information about protein structure with functional and phenotypic information associated with known variants from multiple sodium channelopathies. We then detail the quantitative epileptiform and background EEG features associated with GoF and LoF variant categories, compare the features to neurophysiological metrics from age-matched neurotypical children, and test for associations with clinical outcomes. We hypothesize that quantitative EEG analysis could be useful in characterizing *SCN2A*-DEE and provide both diagnostic and prognostic value.

## Material and methods

### Subjects

#### *SCN2A* cohort

Longitudinal 10-20 EEG and clinical data from individuals with *SCN2A* variants causing DEE were collected by the Ciitizen^®^ platform. Medical records and EEG recordings were collected from institutions within the United States where consented participants received care using the right of access granted by the Health Insurance Portability and Accountability Act (HIPAA).

Each document was evaluated for relevant data variables, including clinical phenotypes, medication use, therapeutic procedures, hospitalizations, attainment of developmental milestones, and seizure frequency. Extracted data were mapped to a coded clinical concept sourced from standard terminology and underwent review and source document verification by a team of advanced practice providers. Caregivers and/or legal guardians of participants provided broad consent to share de-identified data for research purposes. The generation and subsequent analysis of participant data received determinations of exemption through a central IRB. All reported subject IDs were assigned arbitrarily and are not identifiable.

#### Control cohort

Normative data was selected from an EEG database of over 70,000 subjects (Beacon Clinico- EEG Database™). Potential control recordings were identified by programmatically parsing EEG report text for either evidence of normal findings or lack of evidence of epileptiform discharges. Recordings from subjects with normal findings in all EEG reports were included in the control set. Recordings with a lack of evidence of epileptiform discharges (no mention of “discharge”, “spike”, “IED”, “epilepsy”, or other terms indicating epileptiform activity) were screened using a high-sensitivity version of Beacon’s interictal epileptiform discharge (IED) detection algorithm. Recordings flagged as potentially containing IEDs were reviewed by a board-certified neurologist. Recordings were added to the control set if IEDs were not identified algorithmically, or if a board-certified neurologist verified the absence of IEDs. Control recordings were then filtered to match the *SCN2A* recordings’ ages and durations. Age-matching was conducted via the following constraints to accommodate for the faster rate of EEG changes at young ages: *SCN2A* recordings at ages 0-30 days were matched within one week, recordings at 1 month-2 years were matched within one month, and recordings at over 2 years were matched within 6 months. The final control recording dataset was then selected by filtering the age-matched recordings to those that best matched the corresponding *SCN2A* recording duration, to account for differences in EEG patterns expected during routine EEG vs long-term EEG monitoring. *SCN2A* recordings were matched by duration via the following constraints: *SCN2A* recordings of duration 0-1 hour were matched within 30 minutes, recordings of 1-4 hours were matched to a minimum duration of 30 minutes and within 2 hours, recordings of 4-12 hours were matched to a minimum duration of 2 hours and within 6 hours, and recordings longer than 12 hours were matched to a duration greater than 6 hours. In the case that there were no matches based on duration, the age-matched recording with the most similar duration was selected. A total of 1704 recordings from 1230 subjects were matched to the *SCN2A* cohort recordings, and the resulting control dataset contained at least one matched recording for each of the *SCN2A* recordings.

### EEG analysis

#### Seizures and interictal epileptiform discharges

Seizures were detected using Beacon’s automated seizure detection algorithm (SeizureML™), a unified classifier model that characterizes a spectrum of epileptiform patterns. Recordings with likely seizures were manually labeled by a board-certified epileptologist or certified EEG technologist via the Beacon portal. IEDs were detected using Beacon’s automated IED detection algorithm (SpikeML™), a residual neural network model that has been trained and evaluated on EEG segments labeled by 8 epileptologists, using an approach similar to that described in Jing *et al.* 2020^19^. The algorithm returns 1-second windows containing IEDs, computed in steps of 100ms, with overlapping windows merged to identify periods containing IEDs. Outputs were summarized in 20s non-overlapping windows as the percent of each window containing IEDs, or IED burden, to account for the case where a single detection window may contain multiple IEDs. Periods containing flat, non-biological signal (mean variance < 1 µV^2) were excluded from IED burden computations.

#### Background spectral features

Seizures, IED detection windows, and periods with flat, non-biological signal (mean variance < 1 µV^2) were excluded from background spectral analysis. Recordings were downsampled to 128Hz, filtered with 0.5Hz high pass and 60Hz notch filters, and re-referenced to an average montage. Periods on individual channels with high frequency noise were excluded from analysis (mean line-length > 50 µV, computed in 1s windows, where line-length is defined as (v(n) – v(n-1)) / (t(n) - t(n-1)) * sample rate). Multitaper periodograms were computed for 2-second windows, and averaged across all channels, as well as left channels, right channels, anterior, and posterior channels. A minimum duration of 10 minutes of usable recording was required for relative power to be calculated for a given recording. The minimum duration was required for 9 or 4 channels for the power calculated across all channels, and within specific regions, respectively. Relative power was computed for each periodogram by measuring average power within each of four frequency bands (delta = 1-4Hz, theta=4-8Hz, alpha=8-13Hz, beta=13-30Hz) by normalizing by the sum of the four band-wise powers (e.g., a / (d + t + a + b)). Absolute left-right (LR) asymmetry was defined as the absolute value of the difference between average spectral power over the left and right channels divided by their sum (e.g., |(L-R)/(L+R)|). Anterior-posterior (AP) asymmetry was defined as the average power across anterior channels minus the average power across posterior channels, divided by their sum (e.g., (A-P)/(A+P)).

### *SCN2A* variant characterization

#### Subject variant pathogenicity assessment via consensus approach

A consensus variant classification framework was applied to evaluate the pathogenicity of each missense variant exclusively based on the genetic information of the identified variants. This approach is based on the criteria of the American College for Medical Genetics and Genomics (ACMG) variant classification guidelines with some modifications^20^ (for an overview, see **Supplemental Fig. 1**). This framework includes the following criteria: First, recently developed *in-silico* variant pathogenicity prediction scores were integrated, specifically CADD^21^, Paraz- score,^22^ Missense intolerance ratio (MTR),^23^ and EVE.^24^ Second, an *in-silico* pathogenicity prediction score was included; this score was recently developed specifically to assess the pathogenicity of variants in voltage-gated sodium channels.^25^ Third, aligned with the ACMG guidelines, variant location was evaluated with respect to mutational hotspot on protein sequence and structure using the pathogenic enriched region (PER)^26^ or Essential3D site^27^ annotations, respectively. Fourth, the presence or absence of population variants gathered from gnomAD^28^ or UKBioBank^29^ was considered evidence for benign or pathogenic variant classification. Vice versa, presents of pathogenic classified variants from ClinVar^30^, HGMD^31^, or those previously annotated in expert-curated datasets^32,33^ was considered as evidence for pathogenicity. Fifth, the pathogenicity of a variant was manually assessed based on its location in the 3D Na_V_1.2 protein structure (PDB ID: 6J8E). A variant situated in a confirmed pathogenic variant-enriched protein domain^34^ or any other established disease-associated motif was considered evidence for variant pathogenicity. Conversely, a variant located in a domain known to be enriched for population variants^34^ or situated in flexible regions of the protein provides benign evidence. Lastly, in alignment with the ACMG guidelines, significant functional changes of a variant were assessed in an established electrophysiological readout (for details see ^34,35^) as evidence for pathogenicity. A lack of functional change was considered evidence that the variant is benign.

#### Classification of presumed subject variant function

The primary objective of the second layer in variant classification was to differentiate between variants that likely result in a molecular GoF or a LoF of voltage-gated sodium channel function. In alignment with previous work,^32,36,37^ a consensus framework was established that integrated five evidence criteria for the function classification of each variant (for an overview, see **Supplemental Fig. 2**). As a first criterion, two previously developed voltage-gated sodium channel variant function prediction scores were applied.^25,38^ As second criterion, variant proximity to protein structure regions with variants of homogeneous molecular readouts (e.g. either GoF or LoF) was considered as evidence for the functional effect (see **Figure 1**, and Iqbal et al., 2022^27^). For example, a variant that is located within a 3D protein structure region where all other previously reported variants showed a GoF effect was also considered to likely cause a GoF effect. Variants in voltage-gated sodium channels have strong genotype- to phenotype to molecular function concordance.^36^ Therefore, as a third criterion, the local enrichment of variants associated with phenotypes was calculated as a proxy for variant function, and variant scoring was performed similarly to the second criterion. Specifically, *SCN1A* Dravet Syndrome,^39^ *SCN2A* Autism,^33^ and *SCN8A* neurodevelopmental disorders without epilepsy in SCN8A^37^ were considered phenotypes caused by LoF variants. Conversely, variants leading to early onset (<3 months) developmental epilepsy (DEE) in *SCN1A,*^40^ *SCN2A,*^32^ and *SCN8A*^37^ were grouped as those likely leading to a molecular GoF effect. As a fourth criterion, information on Na_V_1.2 domain function identified in previous structure-to-function analyses was included as additional classification evidence.^34^ Fifth, electrophysiological readouts of previously tested variants^34,35^ were mapped to the set of missense variants from this *SCN2A* cohort. If these readouts were interpreted by the authors as definitive GoF or LoF effects across all tested splice forms, it was considered strong evidence for the functional effect. The combination of criteria was scored as a continuum from definite LoF (5) to definite GoF (-5), where the score reflected the level of certainty in the classification (i.e. 0 indicates an uncertain function effect, see Supplemental Fig. 2). Frameshift and termination variants, which result in truncation, were presumed to confer LoF. For statistical modeling, the continuous scores were mapped to “GoF” (-1, -2, -3, -4, -5) and “LoF” (1, 2, 3, 4, 5), and four subjects with variants of uncertain functional effect (0) were excluded from all analyses.

**Figure 1.**
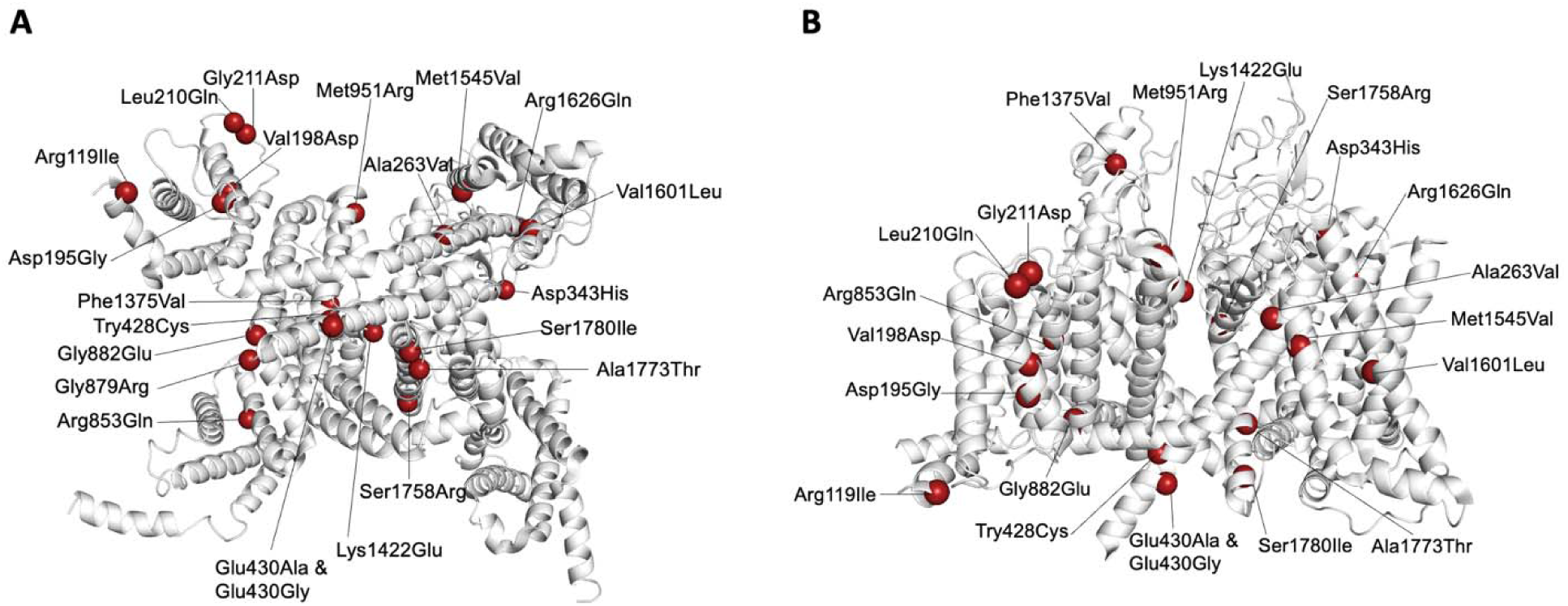
*SCN2A* cohort variants visualized on *SCN2A* 3D protein structure (PDB-ID: 6J8E). (A) Bottom view of missense variants visualized on protein structure (B) Side view of missense variants visualized on protein structure.

#### Protein structure variant mapping

The protein structure (PDB-ID: 6j8e) was taken from the Protein Data Bank.^41^ Missense variants were visualized on the protein structure using Pymol^42^ software.

### Metadata featurization

#### Medication status

Medications were aggregated from medical records by Ciitizen^®^ and restricted to anti-seizure medications (ASMs). The following medications were included in the ASM count: ACTH, brivaracetam, cannabidiol, carbamazepine, clobazam, clonazepam, clorazepate, epidiolex, ethosuximide, felbamate, gabapentin, lacosamide, lamotrigine, levetiracetam, methylprednisolone, oxcarbazepine, perampanel, phenobarbital, phenytoin, prednisolone, prednisone, rufinamide, stiripentol, tetrahydrocannabinol, topiramate, valproate, vigabatrin, zonisamide. The rescue medications diazepam, fosphenytoin, lorazepam, and midazolam were excluded from the ASM count. A subject was considered to be on a sodium channel blocker if they were taking the following ASMs: carbamazepine, lacosamide, lamotrigine, oxcarbazepine, and phenytoin.

ASMs extracted from medical records underwent a process of de-duplication where like data were condensed into date ranges containing a start and end date. ASMs were considered to be in active use from the start date to the end date, inclusive. ASMs whose start and end date were identical were included in the count of active medications only for that single day. When determining the overall number of active ASMs on any given day, only distinctly named medications were considered (i.e., overlapping administrations of a common ingredient were counted a single time). The maximum concurrent ASM count was computed within bins of one month for each subject, to summarize the number of ASMs each subject was on. Across the one month period, the maximum number of active ASMs may be reached more than once for distinct ASM combinations; in this scenario, only the chronologically latest instance was utilized for determining the set of names in this largest concurrent ASM grouping.

#### Clinical history

Reported seizure history and diagnoses were aggregated from medical records by Ciitizen^®^. Reported seizure rates were computed from reported seizure counts, excluding myoclonic seizures, infantile spasms, and absence seizures as they are difficult to detect without EEG and reported counts are therefore unreliable. Subjects with any reported diagnosis of “Autism spectrum disorder” or “Autistic features” were considered to have an autism diagnosis.

#### Reported gross motor developmental delay

Gross motor milestones were the most frequently reported milestone class within the *SCN2A* cohort. For each milestone (e.g., ability to walk), entries indicating that subjects were unable to meet that milestone were compared to a published expected normal age range for that milestone (**Supplementary Table 2**). If the age associated with the “unable” entry was older than the normative range, the entry was considered to be a reported gross motor delay. Recordings prior to a subject’s first reported gross motor delay were assigned a status of “no delay.” Recordings within 6 months after a reported gross motor delay were assigned a status of “delay.” Recordings from subjects without reported gross motor milestones, or that occurred after 6 months of any reported gross motor delay, were not assigned a delay status. A total of 310 EEG recordings from 18 subjects were assigned a gross motor developmental status using this strategy based on age proximity.

### Statistical modeling

Mixed effects logistic regression models were used to estimate the effect of EEG biomarkers on reported gross motor developmental delay. All models underwent a backward selection process wherein the more complex model was preferred when a likelihood ratio test was significant at the 0.05 level. The selection path first simplified random effects, then fixed effects. Note however that the by-subject random intercept was never dropped, to unequivocally account for systematic differences between subjects. Fixed effect 95% confidence intervals were computed for the selected model based on covariance matrices.

Gross motor developmental delay status at the time of recording was used as a binary, categorical, dependent variable in mixed effects logistic regression models. The milestones used to assess normal developmental progress are naturally associated with particular ages, and thus developmental delay is defined, at least in part, by age. As such, age was excluded from all models. To account for age-related changes in background spectral features, age-matched control values for each EEG feature were used as a reference. Deviations from these control values were then used as independent variables in the model rather than the EEG feature values directly (described below). IED burden was not referenced to control values since controls had no or few IEDs at any age. EEG feature predictors were centered for numerical stability. Variant classification was included as a binary, categorical, effects-coded predictor. The maximal model included the following predictors: 95^th^ percentile IED burden or relative spectral power deviation from control, variant classification, interaction of 95^th^ percentile IED burden and variant classification, as well as a subject-level random intercept.

#### Age-Matched Feature Referencing

EEG features computed from the age- and duration-matched control recordings from the Beacon Clinico-EEG Database™ were used as a reference to define deviation from control. The median and median absolute deviation (MAD) of each EEG feature were computed across all control recordings within each developmental age bin (0-2mo, 2-6mo, 6mo-1yr, 1-2yr, 2-4yr, 4-8yr, 8- 16yr). These were paired with the centers of each age bin, i.e., the mean of a bin’s endpoints. The pairs were then linearly interpolated, providing a piecewise-linear reference that evolves with age. The slopes of the lines outside of the range of bin centers, i.e., less than the youngest bin center or greater than the oldest, were taken to be the same as the slope between the closest two points. The resulting curves were determined by visual inspection to be representative of the underlying patterns in the control recordings. Deviations from control for *SCN2A* recordings were computed as robust z-scores using interpolated control medians and MADs for the age at each *SCN2A* recording. Thus, for a given value computed from an *SCN2A* recording and corresponding control median and MAD, the recording’s deviation from control was computed as (value - median) / MAD.

### Statistical Analysis

The fit of the statistical models was evaluated using receiver-operating characteristic (ROC) curves generated using the fitted probabilities from the model to classify observed values, using a range of classification thresholds. In addition to visually inspecting the ROC curves, the area under the curve (AUC) for each ROC was reviewed to assess model fit. The minimum AUC value across both models and variants (GoF, LoF) was 0.88.

## Results

### *SCN2A* and control cohort demographics

The cohort of 28 *SCN2A* subjects included two distinct predicted variant classifications: gain-of-function (GoF, 11 subjects) and loss-of-function (LoF, 17 subjects) (**Table 1**). 493 EEG recordings across the 28 subjects, ages 1 day to 16 years, were included in at least one analysis.

**Table 1.** *SCN2A* cohort demographics by variant classification. . Subject count, phenotype number, and autism number are raw counts, and all other values shown are median +/- interquartile range across subjects. The counts include all *SCN2A* subjects with an assigned variant classification, epilepsy phenotype, and at least one recording included in an analysis. Recording counts include all recordings included in at least one analysis.

| SCN2A variant classification | Subject count | Phenotype | SCN2A diagnosis age (years) | First reported seizure or epilepsy diagnosis age (years) | Subjects with autism diagnosis | Median number of prescribed ASMs | Median number of prescribed SCBs | Recordings per subject | Median recording age (years) |
| --- | --- | --- | --- | --- | --- | --- | --- | --- | --- |
| **GoF** | 11 | EO: 11 | 0.85 +/- 5.83 | 0.0 +/- 0.01 | 2 | 2.0 +/- 1.0 | 1.0 +/- 0.0 | 16.0 +/- 27.0 | 0.49 +/- 5.26 |
| **LoF** | 17 | EO: 1,<br>LO: 7,<br>LOIS: 9 | 1.97 +/- 2.34 | 1.0 +/- 1.13 | 10 | 1.0 +/- 2.0 | 0.0 +/- 0.0 | 13.0 +/- 16.0 | 4.16 +/- 2.58 |

Normative data was assembled from 1230 children without an epilepsy diagnosis or epileptiform features based on neurologists’ review, resulting in 1704 age- and duration-matched control EEG recordings (ages 0 days to 16 years) included in at least one analysis (**Supplementary Table 1**).

*SCN2A* diagnosis lagged behind onset of seizures, often by almost a year. GoF subjects tended to be on more ASMs overall than their LoF counterparts. GoF subjects were prescribed a median of 1 ASM whose primary mechanism was blockade of sodium channels throughout their medical history, consistent with using a partial blockade to reduce seizures caused by overactive sodium channels. Reported seizure history was available for all but 4 LoF subjects. Reported seizure rates varied throughout patients’ lifetimes **(Supplemental Fig. 3).** GoF subjects reported seizure freedom (ie. a seizure count of zero) a median of 15% of the time they reported seizure count, and LoF subjects reported seizure freedom a median of 19% of the time. For some subjects, reported seizures appeared to remit and then return months or years later, and in other subjects there was no consistent reduction in seizures, highlighting the need for improved treatment and diagnostics.

### *SCN2A* functional characterizations align with expected phenotypic indicators

*SCN2A* subjects can be classified into distinct clinical phenotypes: early onset (EO) and late onset, with EO subjects having their first seizure before three months of age. Late onset subjects can be further divided into those with infantile spasms (LOIS) and without (LO). As expected, *SCN2A* phenotype correlated strongly with variant classification. All 11 GoF subjects exhibited an EO phenotype (**Table 1**). 16 LoF subjects exhibited a late onset phenotype (9 LOIS, 7 LO), and only one LoF subject exhibited an EO phenotype. The LoF subject with an EO phenotype was assigned a variant classification of “Likely LoF”.

Of the 28 subjects, 12 had an autism diagnosis, with the majority of those being LoF subjects as expected (2 GoF, 10 LoF subjects diagnosed with autism). Additionally, almost all subjects reported at least one gross motor developmental delay. 27 of the 28 subjects were matched to a gross motor developmental delay status; of the 24 subjects with at least one reported gross motor developmental delay, there were 10 GoF subjects (median age of first reported delay: 1.30 years) and 14 were LoF subjects (median age of first reported delay: 1.14 years).

The median age for *SCN2A* diagnosis was higher in the LoF group (1.97 years vs 0.85 years for GoF), but there was no statistically significant difference in age at diagnosis due to high variability within both groups (p=0.16, Mann Whitney U Test).

### *SCN2A* LoF subjects’ EEG background is dominated by slow rhythms

In the first years of life, the dominant EEG rhythm increases in frequency, shifting from the delta to theta and finally settling into a posterior-dominant rhythm in the alpha band.^43,44^ Relative power was computed in the delta, alpha, theta, and beta bands for 11 GoF, 15 LoF, and 1206 control subjects (230, 195, and 1670 recordings, respectively). The expected shift in the dominant rhythm can be seen in the controls (**Fig. 2**), as exhibited by the gradual decline in relative delta power with age, increase and plateau of relative theta with age, and increase of relative alpha and beta with age.

**Figure 2.**
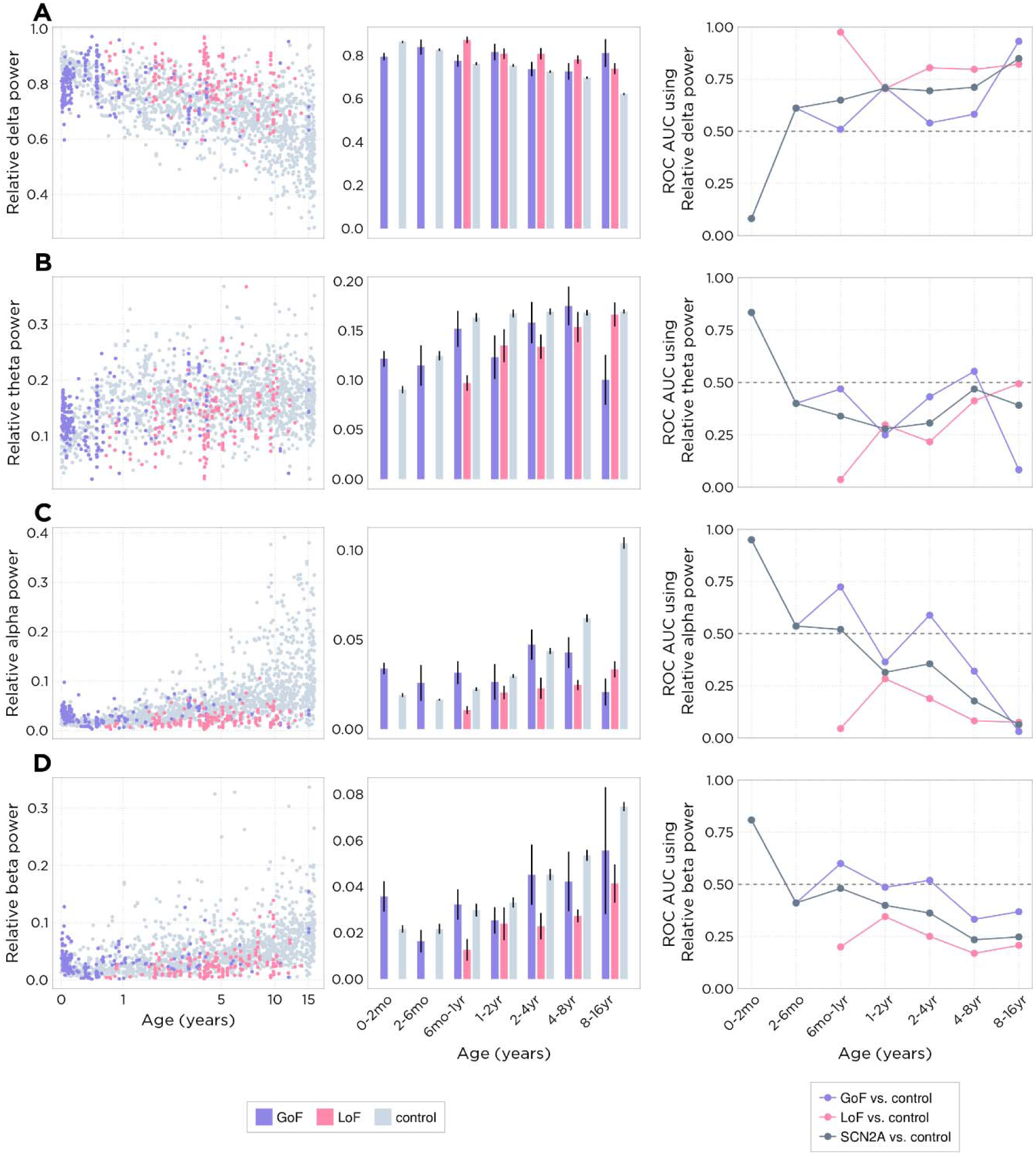
Relative band-wise spectral power by age for *SCN2A* and control subjects. (A-D) Relative delta, theta, alpha, beta power, respectively; Column 1: Per-recording scatter plots; Column 2: Bar plots representing per-subject values within each variant characterization / age bin; Column 3: ROC-AUC curves showing predictive power of each EEG feature. Individual points represent the AUC of an ROC curve computed by classifying subject variant classification using the EEG feature value, sweeping across a range of classification thresholds.

These patterns were less well developed in the *SCN2A* subjects, and most notably so in LoF subjects. Relative delta power was elevated in LoF subjects compared to controls and GoF subjects (**Fig. 2A)**. At younger ages, there was a corresponding decrease in all other relative power values (**Fig. 2B-D**). After 4 years of age, relative theta power matched control values, and the increased relative delta band power appeared to come at the expense of the alpha and beta bands. Elevated relative delta could represent persistent slowing, which would be consistent with delayed development of normal brain rhythms. It could also reflect transient, high-amplitude paroxysmal delta waves that drive up the per-recording median relative delta power value.

Frontal-occipital alpha power asymmetry (FO alpha asymmetry) measures the development of the posterior-dominant rhythm. Frontal-occipital power asymmetry was calculated for 11 GoF, 14 LoF, and 1190 control subjects (201, 166, and 1651 recordings, respectively) (**Fig. 3A**, **Supplemental Fig. 4**). As expected, control subjects’ FO alpha asymmetry values became more negative with age, reflecting the alpha rhythm’s increase in amplitude and localization to posterior channels (**Fig. 3A**). In contrast, FO alpha asymmetry in LoF *SCN2A* subjects remained consistently near zero. This suggests that relative alpha power is not uniformly decreased; instead, the development of the posterior dominant rhythm appears to be impaired in LoF *SCN2A* subjects.

**Figure 3.**
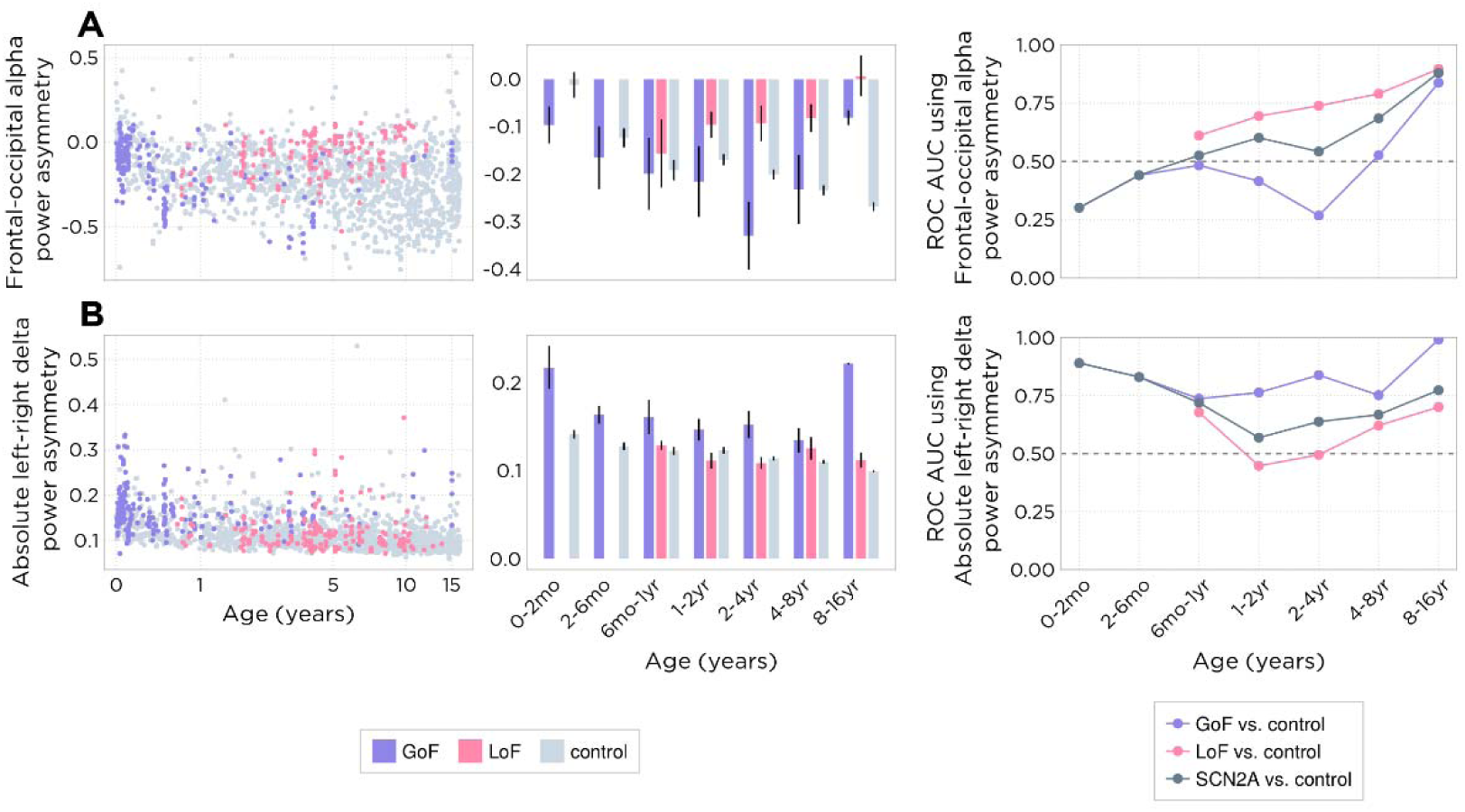
Spectral power asymmetry in *SCN2A* and control subjects. **(A)** frontal-occipital alpha power asymmetry; **(B)** absolute left-right delta power asymmetry; Column 1: Per- recording scatter plots; Column 2: Bar plots representing per-subject values within each variant characterization / age bin; Column 3: ROC-AUC curves showing predictive power of each EEG feature. Individual points represent the AUC of an ROC curve computed by classifying subject variant classification using the EEG feature value, sweeping across a range of classification thresholds.

### GoF *SCN2A* subjects exhibit elevated left-right asymmetry

Absolute left-right delta power asymmetry (LR delta asymmetry) was calculated for 11 GoF, 15 LoF, and 1154 control subjects (228, 195, and 1605 recordings, respectively) (**Fig. 3B**, **Supplemental Fig. 5**). LR delta asymmetry was elevated in GoF subjects compared to controls and LoF subjects across all age groups (**Fig. 3B)**. The degree of elevated asymmetry tended to be modest, and notably below 0.5, the threshold typically used to clinically describe paroxysmal asymmetry. That said, LR delta asymmetry could be used to distinguish GoF subjects from controls relatively well, with AUC values >= 0.74 at any age. In addition to the delta band, elevated asymmetry was also apparent in the theta and alpha bands (**Supplemental Figure 3A- B**), but elevated asymmetry appeared to be most consistent across age in the delta band. This subtle asymmetry may represent focal pathology that can be captured quantitatively but would likely be missed by qualitative visual analysis.

### Frequent IEDs are common across *SCN2A* variants

Reports of IEDs, including multifocal IEDs, are common in *SCN2A* clinical EEG findings^17^. Here, IED prevalence was quantified using Beacon’s automated IED detection algorithm and summarized over 20-second windows to report IED burden throughout each recording. IED burden was calculated for 11 GoF, 17 LoF, and 1226 control subjects (260, 229, and 1696 recordings, respectively). 95^th^ percentile IED burden values, representing a robust maximal IED burden value, are shown for each recording in **Figure 4**. A 95^th^ percentile IED burden value of 0 indicates that IEDs occur rarely, or not at all, within a recording. A 95^th^ percentile IED burden of 50 indicates that the maximal concentration of IEDs within a recording consisted of either intermittent periodic IEDs, or IEDs spaced in time approximately once every 2s.

**Figure 4.**
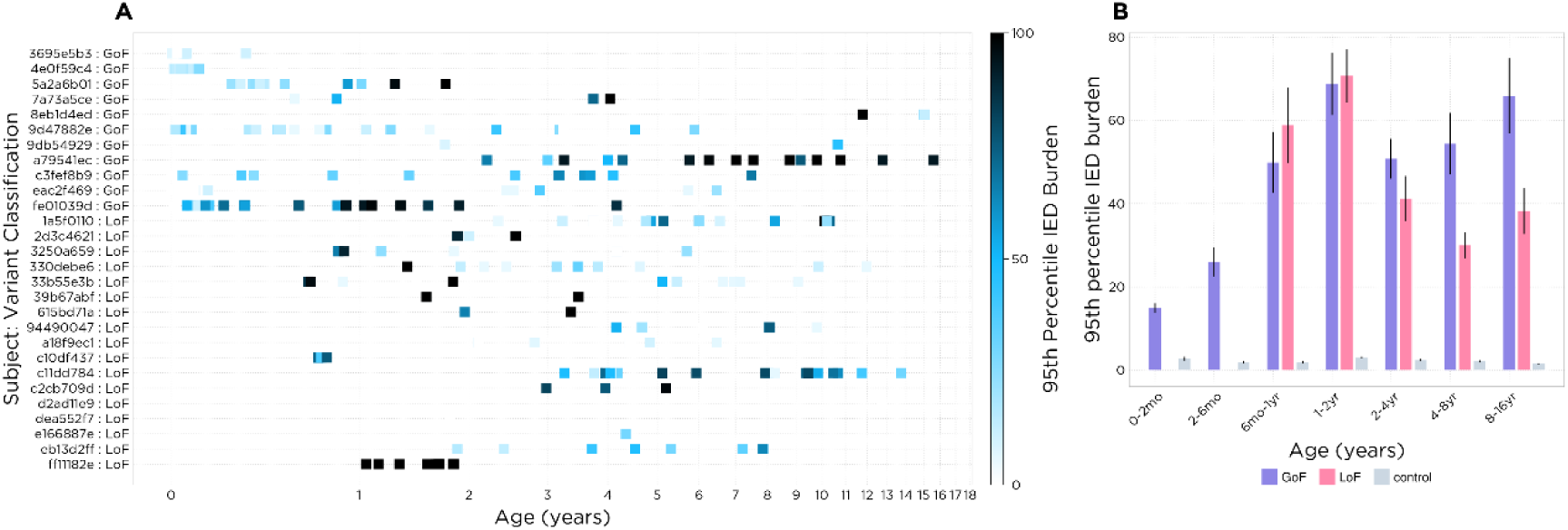
IED burden in *SCN2A*. **(A)** 95th percentile IED burden for each included recording in the *SCN2A* dataset using automated IED detection (SpikeML™); **(B)** 95th percentile IED burden within each variant classification / age range compared to age-matched controls (grey).

Overall, IED burden appeared to increase with age over the first year of life and then remained high (∼50%) in GoF subjects, whereas for LoF subjects IED burden appeared to drop below that of GoF subjects after 2 years of age. As expected, 95^th^ percentile IED burden was low -- approximately 2% -- for control subjects regardless of age. This non-zero value represents occasional IEDs expected to occur in a subset of normal pediatric subjects combined with a non-zero false-positive detection rate. In any case, IED burden was elevated in *SCN2A* subjects, particularly in those with GoF variants.

### High IED burden and spectral abnormalities correlate with reported gross motor developmental delay

The relationship between EEG features and reported gross motor delay was assessed using a set of logistic mixed effects models with reported gross motor developmental delay as the response variable. The effect of IED burden on the probability of reporting a gross motor delay was assessed via a model including 95^th^ percentile IED burden and *SCN2A* variant classification as predictors (7 GoF and 11 LoF subjects, with 197 and 70 recordings, respectively, were included in the model). Note that age was omitted from the set of possible predictors, as the response variable is determined by age. The selected model reported a significant main effect of IED burden (p = 0.0015), variant classification (p = 1e-05), and a significant interaction between IED burden and variant classification (p = 1e-06) (**Table 2, Supplemental Fig. 6**). The model estimated that GoF subjects with higher IED burden were more likely to report a gross motor delay (**Fig. 5A**). In contrast, LoF subjects were always likely to be delayed, which is unsurprising given that the first symptom for these subjects is often developmental delay, followed by epileptic activity. Because EEGs are unlikely to be conducted when the main concern is developmental delay, there were only 14 recordings included in the model from LoF subjects that were made before the first reported developmental delay, making the model unlikely to draw inferences about LoF subjects without reported gross motor delay. In contrast, GoF subjects with higher IED burden were more likely to be developmentally delayed.

**Figure 5.**
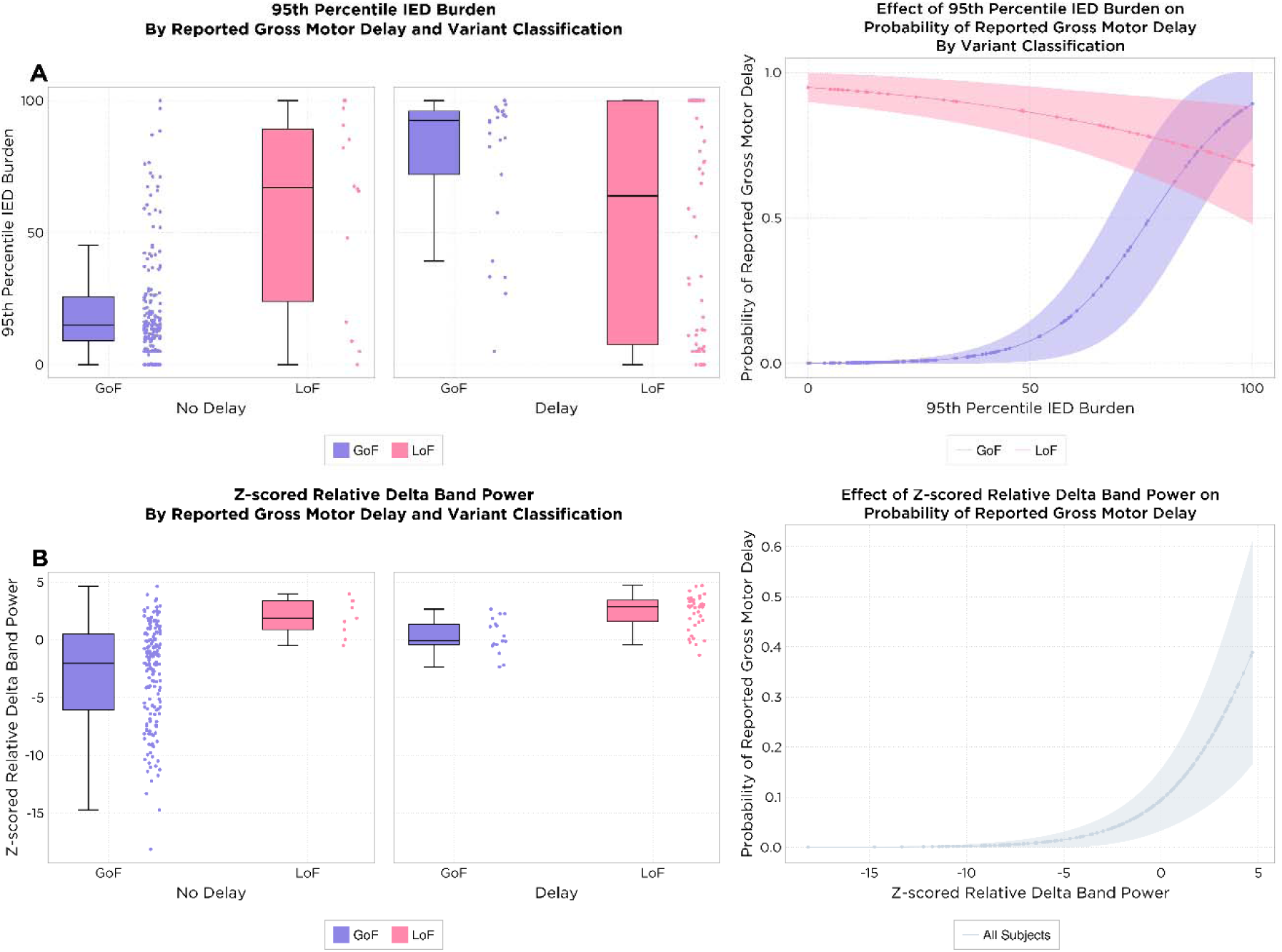
Effect of IED burden and relative delta power on probability of gross motor delay. **(A)** Left: Distribution of 95^th^ percentile IED burden by *SCN2A* variant classification and gross motor developmental delay. Right: Effect of 95th percentile IED burden on gross motor developmental delay for GoF and LoF patients from a mixed effects logistic regression model. Lines represent model predictions and shaded bars represent model uncertainty. **(B)** Left: Z-scored relative delta distribution by *SCN2A* variant classification. Right: Effect of z-scored relative delta on gross motor developmental delay for GoF and LoF patients from a mixed effects logistic regression model.

**Table 2.**
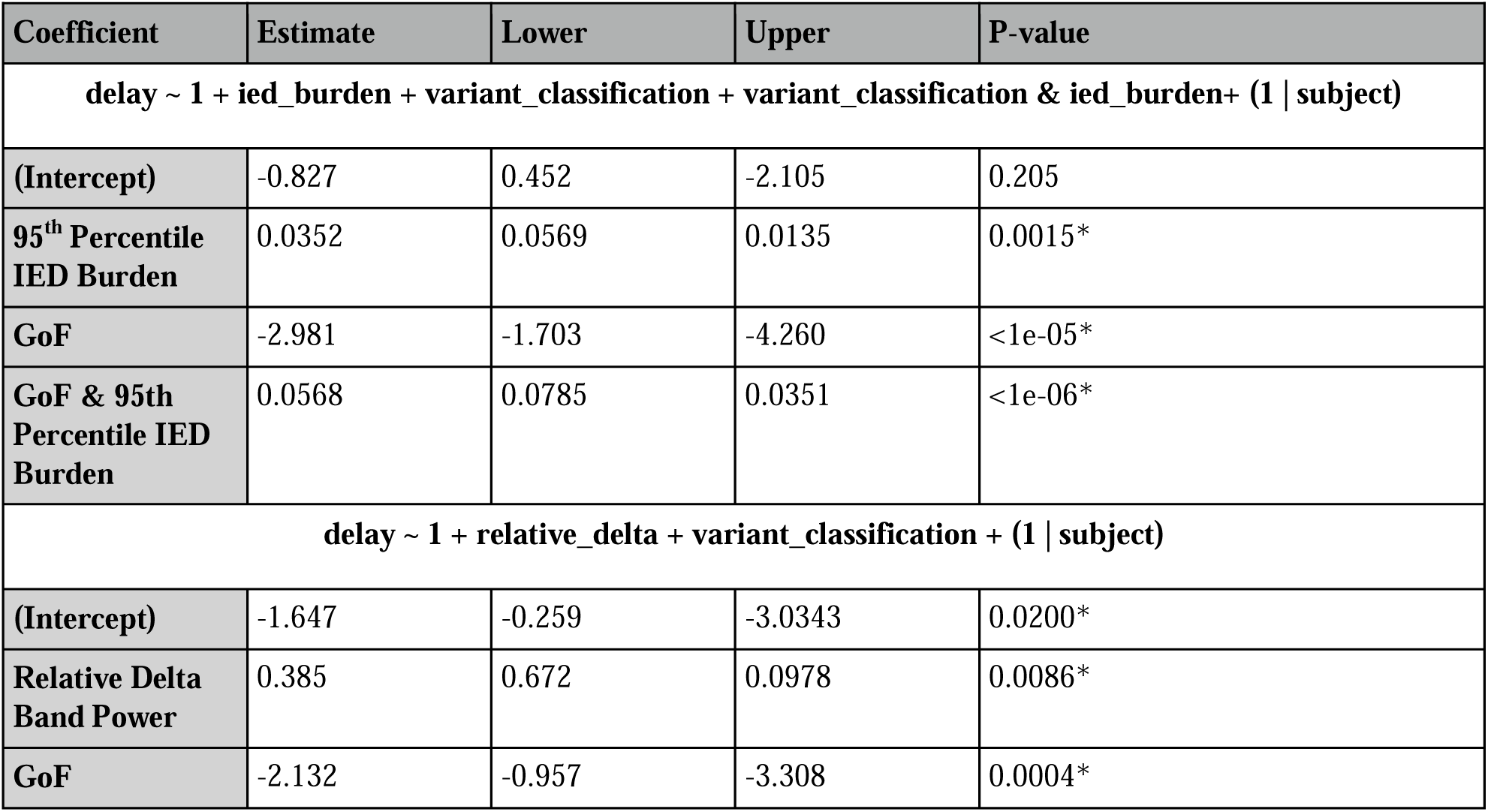
Model coefficients table. Top: Coefficients for model of effect of IED burden on probability of gross motor delay. Bottom: Coefficients for model of effect of relative delta on probability of gross motor delay.

To assess the relationship between spectral features and reported gross motor deficits, z-scores representing deviation of spectral features from age-matched controls were computed. These z-scores account for expected age-related changes in spectral power and allow age to be omitted from the statistical models. A mixed effects model including z-scored relative delta power and *SCN2A* variant classification revealed a positive association between z-scored relative delta power and the probability of reporting gross motor developmental delay (7 GoF and 9 LoF subjects, with 183 and 47 recordings, respectively, were included in the model) (**Fig. 5B**). The model included a significant main effect of z-scored relative delta power (p = 0.0086), and a significant effect of *SCN2A* variant classification (p = 0.0004), but no significant interaction between the two (**Table 2, Supplemental Fig. 7**). The more abnormally elevated a subject’s delta power was with respect to age-matched controls (more positive z-score), the more likely they were to report a gross motor delay, suggesting that the persistence of slower background features correlates with worse clinical outcome in DEE.

## Discussion

In this study we utilized a rich *SCN2A* dataset of 28 children requiring neurological care to evaluate quantitative EEG as a biomarker of developmental delay. In addition to quantifying a range of EEG abnormalities, we demonstrate the link between EEG biomarkers and gross motor developmental delay. In particular, we demonstrate two core findings: background spectral features are correlated with motor developmental outcome when measured relative to age-matched neurotypical children, and interictal epileptiform activity is correlated with delayed motor development in subjects with GoF variants. These findings suggest the importance of EEG as a prognostic tool beyond the standard clinical metrics.

To understand the association between EEG abnormalities and underlying sodium channel function, we employed a novel, genetics-based consensus framework to infer the functional characterization of the *SCN2A* variants in this cohort. This highly scalable approach provides a more granular description of underlying biophysical function, which was leveraged in this study to identify subjects with GoF and LoF mutations rather than relying on phenotypic classification. Future studies could take full advantage of the continuum provided by this consensus framework to identify whether the presence or severity of particular EEG abnormalities correlate with consensus score.

EEG is routinely performed to assess the need for ASM therapy and is often used to gauge disease severity based on visual inspection. This typically involves visual estimation of background frequency content and is typically limited to defining a dichotomy between “abnormal” and “normal” without quantitative measurements. Pathologically elevated delta power has previously been demonstrated to reflect disrupted network organization^45^. To assess the prognostic value of interictal EEG recordings, we quantified the background spectral power content of EEG from children with *SCN2A*-DEE in comparison to age-matched neurotypical controls. Compared to controls, subjects with *SCN2A*-DEE demonstrated altered evolution of expected EEG maturation and pathological patterns of spectral features at almost all ages. GoF subjects also demonstrated focal abnormalities not seen in other cohorts. Elevated delta power compared to age-matched controls was strongly correlated with gross motor developmental delay, regardless of variant function. Taken together, these findings suggest that EEG biomarkers can be used to identify neurological abnormalities that correlate both with *SCN2A* variants and measures of development.

Background interictal epileptiform activity provided greater insight into the dichotomy between GoF and LoF variants. IEDs were commonly observed in both cohorts. However, we identified a clear relationship between IED burden and the presence of delayed gross motor milestones in GoF subjects only. This relationship was absent in subjects with LoF variants. These findings are consistent with the hypothesis that GoF variants result in an epileptic process that contributes to or causes encephalopathy. Future therapies targeting the underlying genetic cause of this channelopathy will be needed to address this hypothesis – assuming that genetic therapies can be administered at an appropriate age early enough to prevent the development of a static and severely encephalopathic network.

While the evidence presented in this study demonstrates a strong link between EEG biomarkers and functional outcomes in *SCN2A*-DEE, certain variables could not be fully controlled. For example, we acknowledge that ASM regimens were highly variable and could potentially have an independent and cumulative negative impact on development, cognition, and EEG background. In general, understanding the cause-effect link between ASMs and disease progression is complicated, and could be an important topic of a future study to better understand DEE progression. Additionally, EEG measurements were collected at variable timepoints at the treating clinicians’ discretion and may not fully capture the natural disease course. However, despite the challenges inherent to retrospective studies using real-world data, well-controlled prospective studies are often infeasible due to the length of time to collect longitudinal data, patient attrition and cost associated.

These findings demonstrate that EEG, a universally available test for DEE populations, contains critical biomarkers of disease severity. Background spectral features and IED burden could aid in patient selection when therapy carries significant risk. Furthermore, in all SCN2A-DEE patients, EEG could serve as a quantitative biomarker of network pathology, with delta power normalized to age-matched control distributions providing an efficacy measure when treating with recurrent therapy such as antisense oligonucleotides. These data-driven tools introduce an evidence-based personalized medicine paradigm for genetic epilepsies.

## Data Availability

The data produced in the present work are proprietary and unable to be shared.

## Acknowledgements

We thank the *SCN2A* Families Foundation for their support of these research programs.

## References

1. Yu FH, Catterall WA. Overview of the voltage-gated sodium channel family. Genome Biol. 2003;4(3):207. doi:10.1186/gb-2003-4-3-207

2. Wallace RH, Wang DW, Singh R, et al. Febrile seizures and generalized epilepsy associated with a mutation in the Na+-channel ß1 subunit gene SCN1B. Nat Genet. 1998;19(4):366–370. doi:10.1038/1252

3. Baulac S, Gourfinkel-An I, Picard F, et al. A Second Locus for Familial Generalized Epilepsy with Febrile Seizures Plus Maps to Chromosome 2q21-q33. Am J Hum Genet. 1999;65(4):1078–1085. doi:10.1086/302593

4. Sugawara T, Tsurubuchi Y, Agarwala KL, et al. A missense mutation of the Na ^+^ channel α _II_ subunit gene *Na* _v_ *1.2* in a patient with febrile and afebrile seizures causes channel dysfunction. Proc Natl Acad Sci. 2001;98(11):6384–6389. doi:10.1073/pnas.111065098

5. Kamiya K, Kaneda M, Sugawara T, et al. A nonsense mutation of the sodium channel gene SCN2A in a patient with intractable epilepsy and mental decline. J Neurosci Off J Soc Neurosci. 2004;24(11):2690–2698. doi:10.1523/JNEUROSCI.3089-03.2004

6. Yamakawa K. Na channel gene mutations in epilepsy--the functional consequences. Epilepsy Res. 2006;70 Suppl 1:S218–222. doi:10.1016/j.eplepsyres.2005.11.025

7. Ogiwara I, Ito K, Sawaishi Y, et al. De novo mutations of voltage-gated sodium channel II gene SCN2A in intractable epilepsies. Neurology. 2009;73(13):1046–1053. doi:10.1212/WNL.0b013e3181b9cebc

8. Ben-Shalom R, Keeshen CM, Berrios KN, An JY, Sanders SJ, Bender KJ. Opposing Effects on NaV1.2 Function Underlie Differences Between SCN2A Variants Observed in Individuals With Autism Spectrum Disorder or Infantile Seizures. Biol Psychiatry. 2017;82(3):224–232. doi:10.1016/j.biopsych.2017.01.009

9. Sanders SJ, Murtha MT, Gupta AR, et al. De novo mutations revealed by whole-exome sequencing are strongly associated with autism. Nature. 2012;485(7397):237-241. doi:10.1038/nature10945

10. Boiko T, Van Wart A, Caldwell JH, Levinson SR, Trimmer JS, Matthews G. Functional Specialization of the Axon Initial Segment by Isoform-Specific Sodium Channel Targeting. J Neurosci. 2003;23(6):2306–2313. doi:10.1523/JNEUROSCI.23-06-02306.2003

11. Gazina EV, Leaw BTW, Richards KL, et al. ‘Neonatal’ Nav1.2 reduces neuronal excitability and affects seizure susceptibility and behaviour. Hum Mol Genet. 2015;24(5):1457–1468. doi:10.1093/hmg/ddu562

12. Hu W, Tian C, Li T, Yang M, Hou H, Shu Y. Distinct contributions of Nav1.6 and Nav1.2 in action potential initiation and backpropagation. Nat Neurosci. 2009;12(8):996–1002. doi:10.1038/nn.2359

13. Wolff M, Johannesen KM, Hedrich UBS, et al. Genetic and phenotypic heterogeneity suggest therapeutic implications in SCN2A-related disorders. Brain. 2017;140(5):1316–1336. doi:10.1093/brain/awx054

14. Berecki G, Howell KB, Heighway J, et al. Functional correlates of clinical phenotype and severity in recurrent SCN2A variants. Commun Biol. 2022;5(1):515. doi:10.1038/s42003-022-03454-1

15. Li M, Jancovski N, Jafar-Nejad P, et al. Antisense oligonucleotide therapy reduces seizures and extends life span in an SCN2A gain-of-function epilepsy model. J Clin Invest. 2021;131(23):e152079. doi:10.1172/JCI152079

16. Kong Y, Yan K, Hu L, et al. Association between SCN1A and SCN2A mutations and clinical/EEG features in Chinese patients from epilepsy or severe seizures. Clin Chim Acta. 2018;483:14–19. doi:10.1016/j.cca.2018.03.027

17. Howell KB, McMahon JM, Carvill GL, et al. SCN2A encephalopathy: A major cause of epilepsy of infancy with migrating focal seizures. Neurology. 2015;85(11):958–966. doi:10.1212/WNL.0000000000001926

18. Brimble E, Kim J, Martin RL, McKnight D, Lacoste AMB. Computation of Longitudinal Phenotypes in 466 Individuals with a Developmental and Epileptic Encephalopathy Enables Clinical Trial Readiness. Genetic and Genomic Medicine; 2023. doi:10.1101/2023.03.02.23286645

19. Jing J, Sun H, Kim JA, et al. Development of Expert-Level Automated Detection of Epileptiform Discharges During Electroencephalogram Interpretation. JAMA Neurol. 2020;77(1):103–108. doi:10.1001/jamaneurol.2019.3485

20. Richards S, Aziz N, Bale S, et al. Standards and guidelines for the interpretation of sequence variants: a joint consensus recommendation of the American College of Medical Genetics and Genomics and the Association for Molecular Pathology. Genet Med Off J Am Coll Med Genet. 2015;17(5):405–424. doi:10.1038/gim.2015.30

21. Rentzsch P, Witten D, Cooper GM, Shendure J, Kircher M. CADD: predicting the deleteriousness of variants throughout the human genome. Nucleic Acids Res. 2019;47(D1):D886–D894. doi:10.1093/nar/gky1016

22. Lal D, May P, Perez-Palma E, et al. Gene family information facilitates variant interpretation and identification of disease-associated genes in neurodevelopmental disorders. Genome Med. 2020;12(1):28. doi:10.1186/s13073-020-00725-6

23. Traynelis J, Silk M, Wang Q, et al. Optimizing genomic medicine in epilepsy through a gene-customized approach to missense variant interpretation. Genome Res. 2017;27(10):1715–1729. doi:10.1101/gr.226589.117

24. Frazer J, Notin P, Dias M, et al. Disease variant prediction with deep generative models of evolutionary data. Nature. 2021;599(7883):91-95. doi:10.1038/s41586-021-04043-8

25. Heyne HO, Baez-Nieto D, Iqbal S, et al. Predicting functional effects of missense variants in voltage-gated sodium and calcium channels. Sci Transl Med. 2020;12(556). doi:10.1126/scitranslmed.aay6848

26. Pérez-Palma E, May P, Iqbal S, et al. Identification of pathogenic variant enriched regions across genes and gene families. Genome Res. 2020;30(1):62–71. doi:10.1101/gr.252601.119

27. Iqbal S, Brünger T, Pérez-Palma E, et al. Delineation of functionally essential protein regions for 242 neurodevelopmental disorders. Brain J Neurol. Published online October 18, 2022:awac381. doi:10.1093/brain/awac381

28. Karczewski KJ, Francioli LC, Tiao G, et al. The mutational constraint spectrum quantified from variation in 141,456 humans. Nature. 2020;581(7809):434-443. doi:10.1038/s41586-020-2308-7

29. Sudlow C, Gallacher J, Allen N, et al. UK biobank: an open access resource for identifying the causes of a wide range of complex diseases of middle and old age. PLoS Med. 2015;12(3):e1001779. doi:10.1371/journal.pmed.1001779

30. Landrum MJ, Lee JM, Benson M, et al. ClinVar: improving access to variant interpretations and supporting evidence. Nucleic Acids Res. 2018;46(D1):D1062–D1067. doi:10.1093/nar/gkx1153

31. Stenson PD, Ball EV, Mort M, et al. Human Gene Mutation Database (HGMD): 2003 update. Hum Mutat. 2003;21(6):577–581. doi:10.1002/humu.10212

32. Wolff M, Johannesen KM, Hedrich UBS, et al. Genetic and phenotypic heterogeneity suggest therapeutic implications in SCN2A-related disorders. Brain J Neurol. 2017;140(5):1316–1336. doi:10.1093/brain/awx054

33. Crawford K, Xian J, Helbig KL, et al. Computational analysis of 10,860 phenotypic annotations in individuals with SCN2A-related disorders. Genet Med Off J Am Coll Med Genet. 2021;23(7):1263–1272. doi:10.1038/s41436-021-01120-1

34. Brunklaus A, Feng T, Brünger T, et al. Gene variant effects across sodium channelopathies predict function and guide precision therapy. Brain J Neurol. Published online January 17, 2022:awac006. doi:10.1093/brain/awac006

35. Thompson CH, Potet F, Abramova TV, et al. Epilepsy-associated SCN2A (NaV1.2) Variants Exhibit Diverse and Complex Functional Properties. Published online February 23, 2023:2023.02.23.529757. doi:10.1101/2023.02.23.529757

36. Brunklaus A, Du J, Steckler F, et al. Biological concepts in human sodium channel epilepsies and their relevance in clinical practice. Epilepsia. 2020;61(3):387–399. doi:10.1111/epi.16438

37. Johannesen KM, Liu Y, Koko M, et al. Genotype-phenotype correlations in SCN8A-related disorders reveal prognostic and therapeutic implications. Brain J Neurol. 2022;145(9):2991–3009. doi:10.1093/brain/awab321

38. Boßelmann CM, Hedrich UBS, Lerche H, Pfeifer N. Learning with phenotypic similarity improves the prediction of functional effects of missense variants in voltage-gated sodium channels. Published online September 30, 2022:2022.09.29.510111. doi:10.1101/2022.09.29.510111

39. Brunklaus A, Pérez-Palma E, Ghanty I, et al. Development and Validation of a Prediction Model for Early Diagnosis of SCN1A-Related Epilepsies. Neurology. 2022;98(11):e1163–e1174. doi:10.1212/WNL.0000000000200028

40. Brunklaus A, Brünger T, Feng T, et al. The gain of function SCN1A disorder spectrum: novel epilepsy phenotypes and therapeutic implications. Brain J Neurol. Published online June 13, 2022:awac210. doi:10.1093/brain/awac210

41. Berman HM, Westbrook J, Feng Z, et al. The Protein Data Bank. Nucleic Acids Res. 2000;28(1):235–242. doi:10.1093/nar/28.1.235

42. Schrödinger, LLC. The PyMOL Molecular Graphics System.

43. Cellier D, Riddle J, Petersen I, Hwang K. The development of theta and alpha neural oscillations from ages 3 to 24 years. Dev Cogn Neurosci. 2021;50:100969. doi:10.1016/j.dcn.2021.100969

44. Marshall PJ, Bar-Haim Y, Fox NA. Development of the EEG from 5 months to 4 years of age. Clin Neurophysiol. 2002;113(8):1199–1208. doi:10.1016/S1388-2457(02)00163-3

45. Houtman SJ, Lammertse HCA, van Berkel AA, et al. STXBP1 Syndrome Is Characterized by Inhibition-Dominated Dynamics of Resting-State EEG. Front Physiol. 2021;12. Accessed July 7, 2023. https://www.frontiersin.org/articles/10.3389/fphys.2021.775172

46. 46. WHO MULTICENTRE GROWTH REFERENCE STUDY GROUP, Onis M. WHO Motor Development Study: Windows of achievement for six gross motor development milestones: Windows of achievement for motor milestones. Acta Paediatr. 2007;95:86–95. doi:10.1111/j.1651-2227.2006.tb02379.x

47. Dosman CF, Andrews D, Goulden KJ. Evidence-based milestone ages as a framework for developmental surveillance. Paediatr Child Health. 2012;17(10):561–568. doi:10.1093/pch/17.10.561

48. Adolph KE, Berger SE, Leo AJ. Developmental continuity? Crawling, cruising, and walking: Developmental continuity. Dev Sci. 2011;14(2):306–318. doi:10.1111/j.1467-7687.2010.00981.x

