## Supplement for "Electroencephalographic insights into variant function and clinical outcomes in *SCN2A* encephalopathy"

**Supplemental Figures**

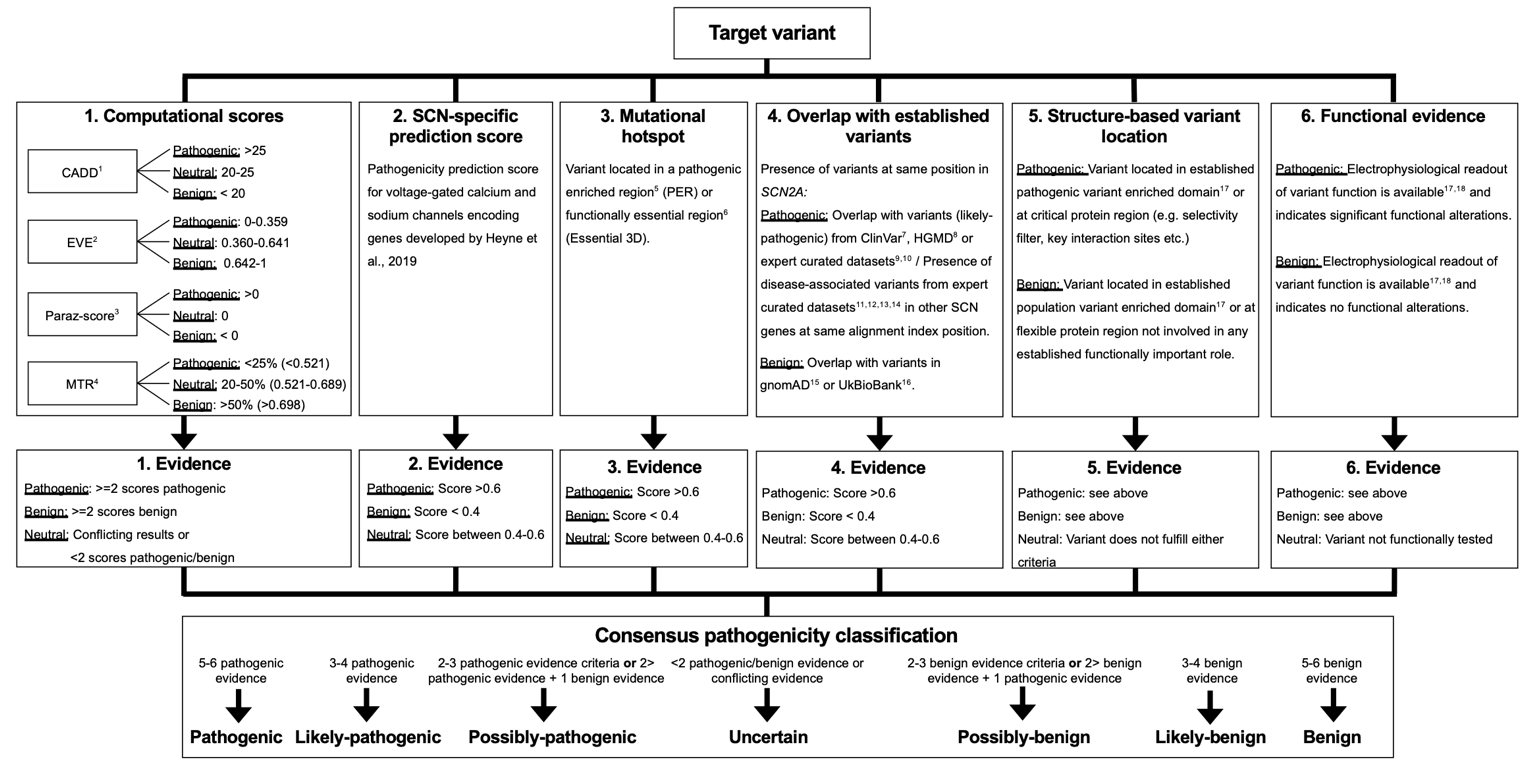

**Supplemental Figure 1**. **Consensus pathogenicity classification based on genetic data.** Variant classification criteria 1-6 were each assigned a score of 1 or -1 for pathogenic or benign respectively. All evidence was then combined into a consensus score ranging from -6 (benign) to 6 (pathogenic) (see Methods for details).

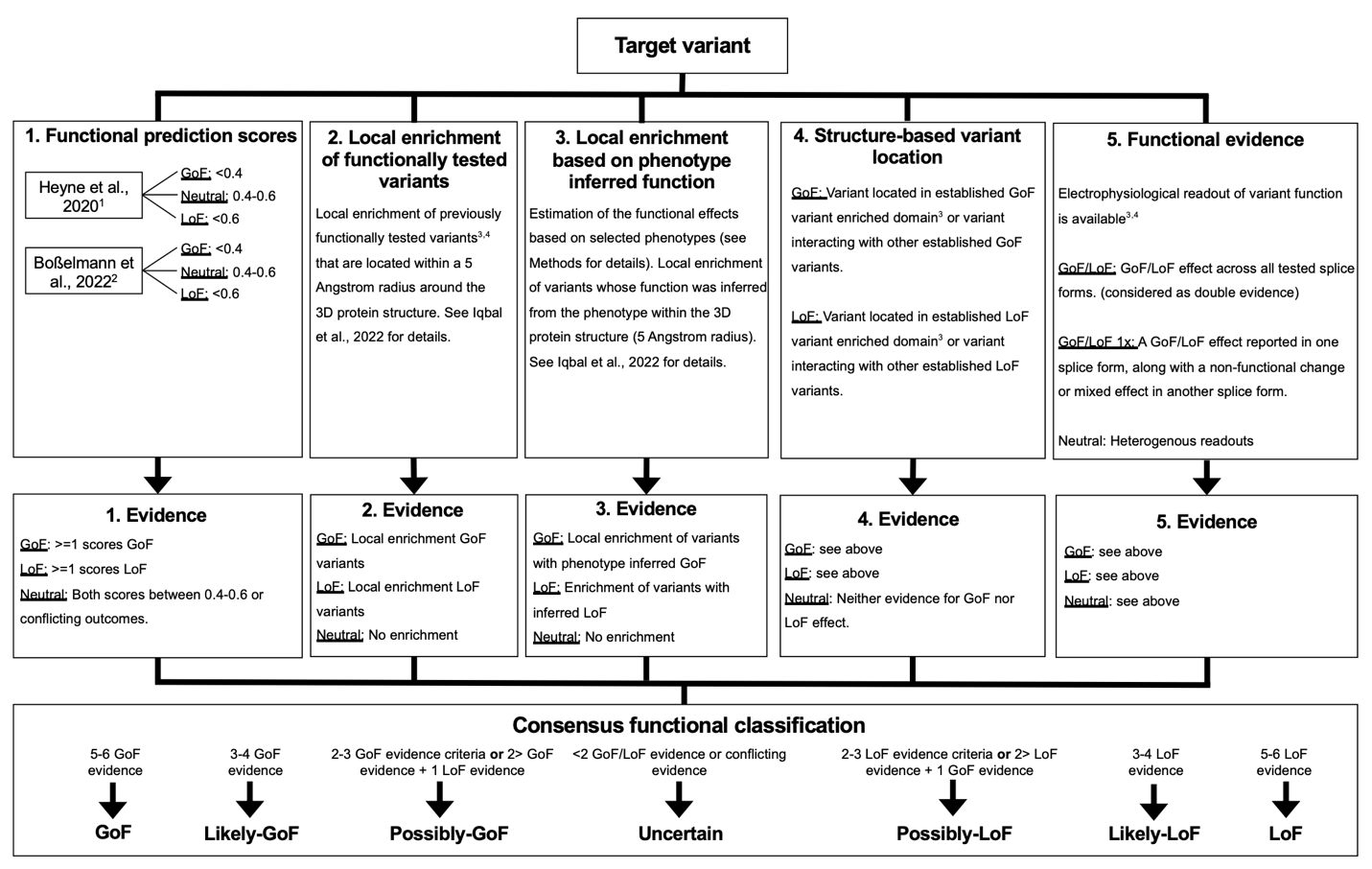

**Supplemental Figure 2.** Consensus functional classification based on genetic data. Variant classification criteria 1-5 were each assigned a score of 1 or -1 for GoF or LoF respectively. Homogenous functional evidence across different isoforms was considered as the only criterium assigning two evidence points. All evidence was then combined into a consensus score ranging from -6 (LoF) to 6 (GoF) (see Methods for details).

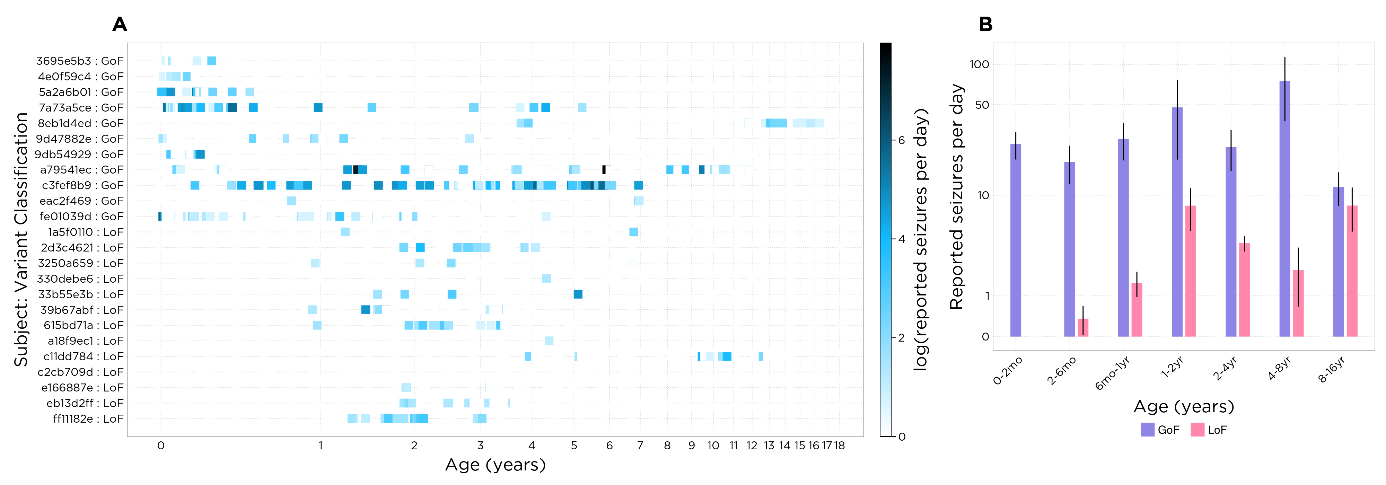
**Supplemental Figure 3. Reported seizures in *SCN2A* subjects.** (A) Seizure rate for each included *SCN2A* subject with reported seizure entries. Seizure rate is displayed as seizures per day on a log scale. (B) Reported seizure rate within each variant classification / age range. Seizure rate is displayed as seizures per day, on a log scale.

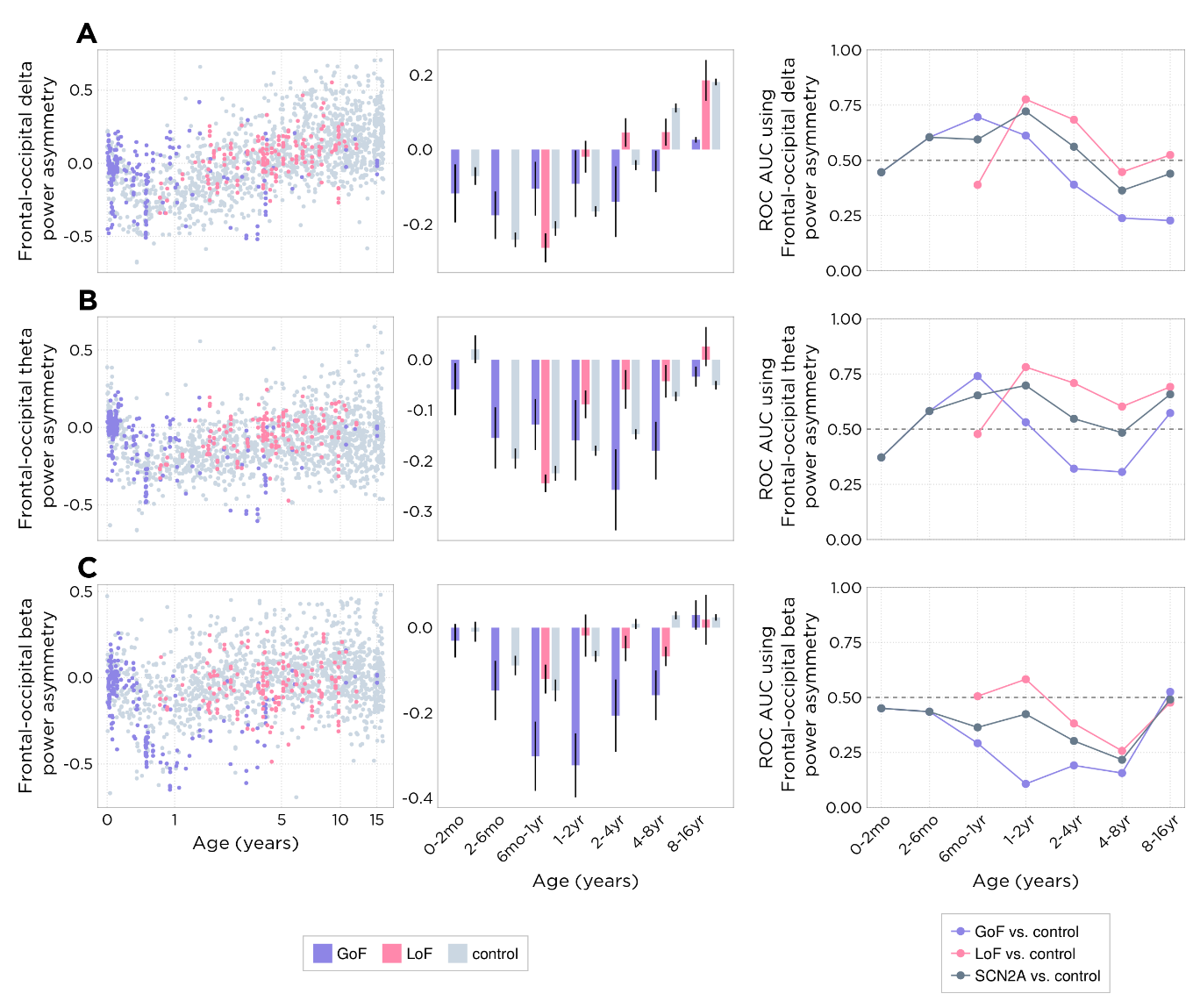

**Supplemental Figure 4.** **Frontal-occipital spectral power asymmetry in *SCN2A* and control subjects. (A-C)** frontal-occipital power asymmetry for delta, theta, and beta bands, respectively; Column 1: Per-recording scatter plots; Column 2: Bar plots representing per-subject values within each variant characterization / age bin; Column 3: ROC-AUC curves showing predictive power of each EEG feature. Individual points represent the AUC of an ROC curve computed by classifying subject variant classification using the EEG feature value, sweeping across a range of classification thresholds.

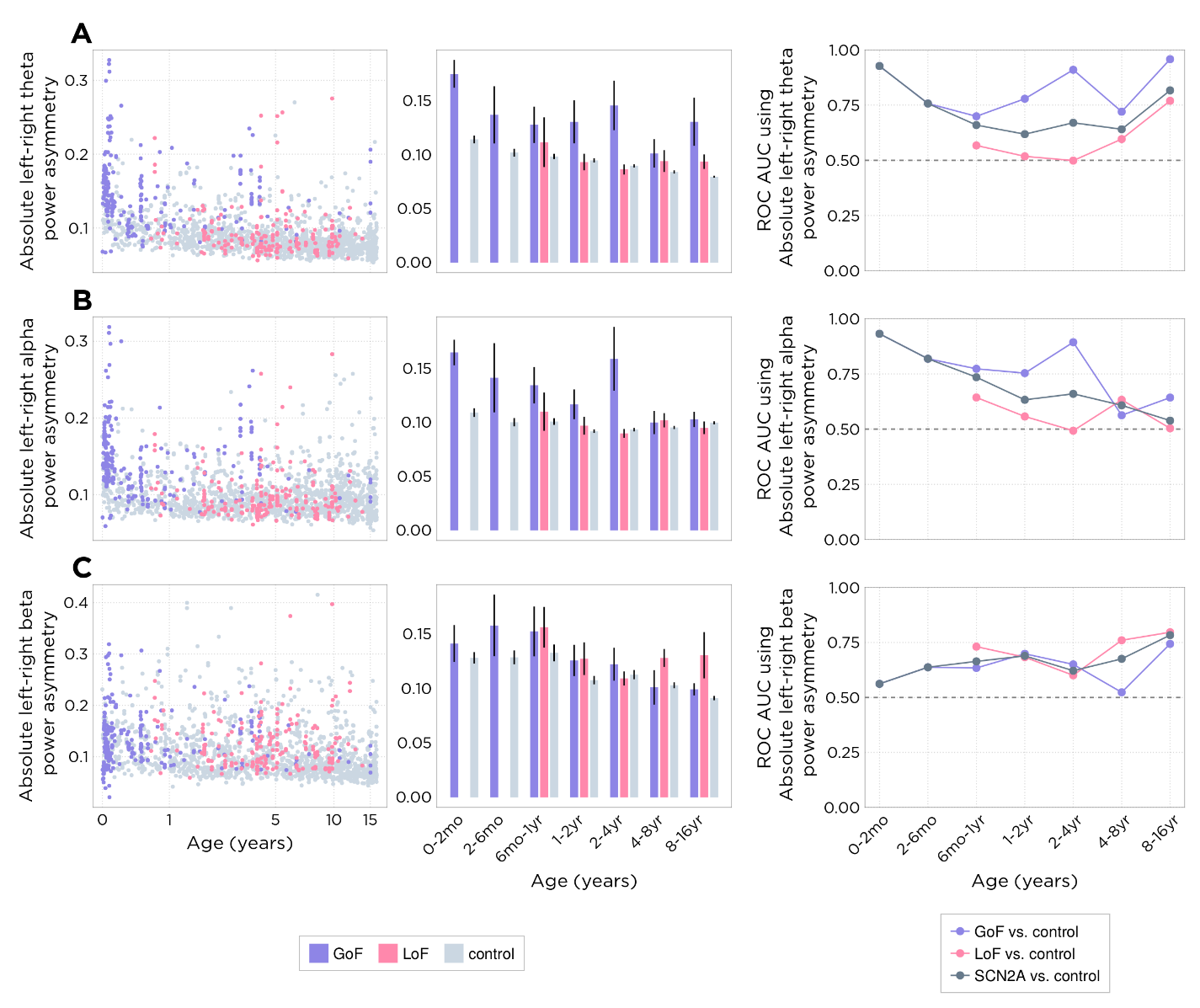

**Supplemental Figure 5. Absolute left-right spectral power asymmetry in *SCN2A* and control subjects. (A-C)** absolute left-right power asymmetry for theta, alpha, and beta bands, respectively; Column 1: Per-recording scatter plots; Column 2: Bar plots representing per-subject values within each variant characterization / age bin; Column 3: ROC-AUC curves showing predictive power of each EEG feature. Individual points represent the AUC of an ROC curve computed by classifying subject variant classification using the EEG feature value, sweeping across a range of classification thresholds.

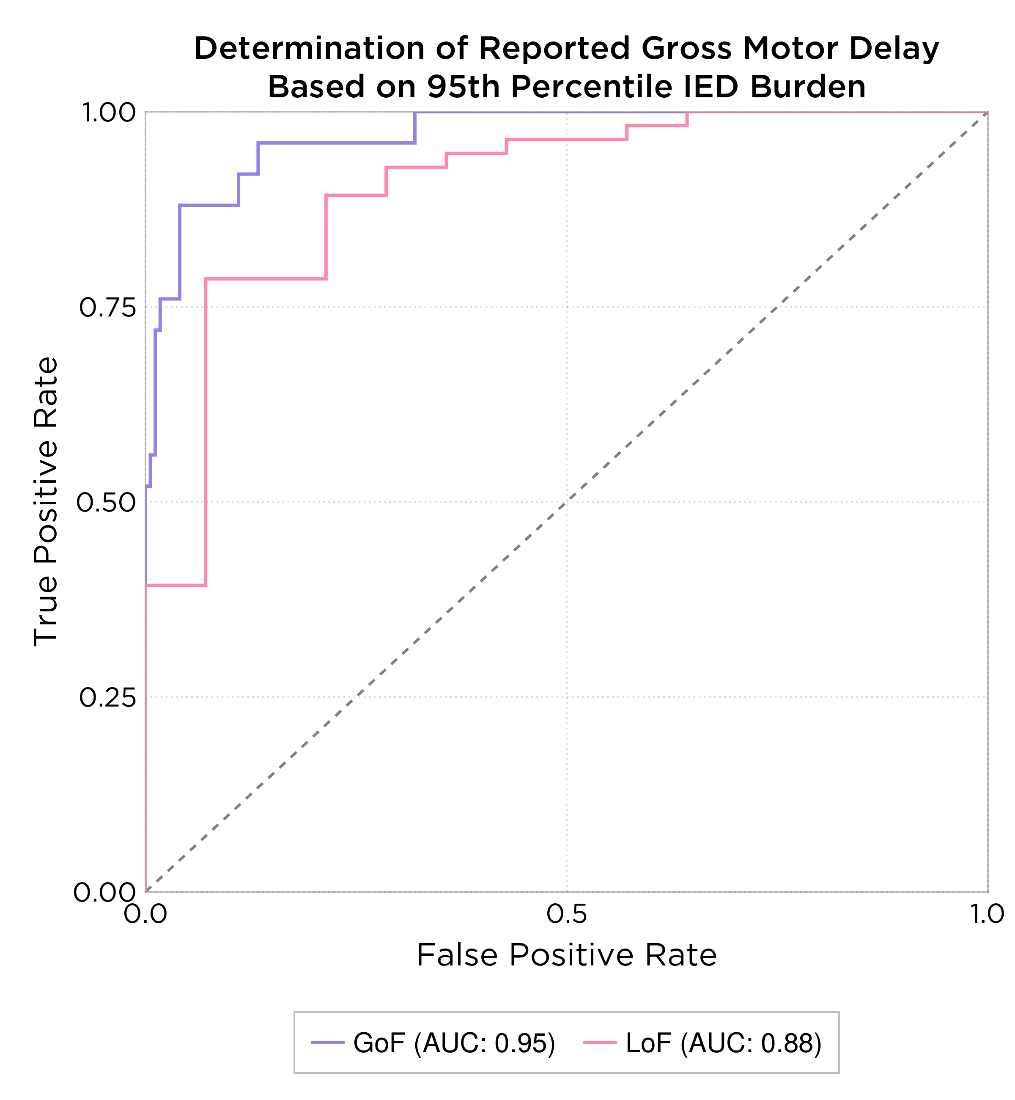

**Supplemental Figure 6. Model fit for model of gross motor developmental delay by IED burden.** ROC curve for model, split by variant classification. GoF AUC: 0.95; LoF AUC: 0.88.

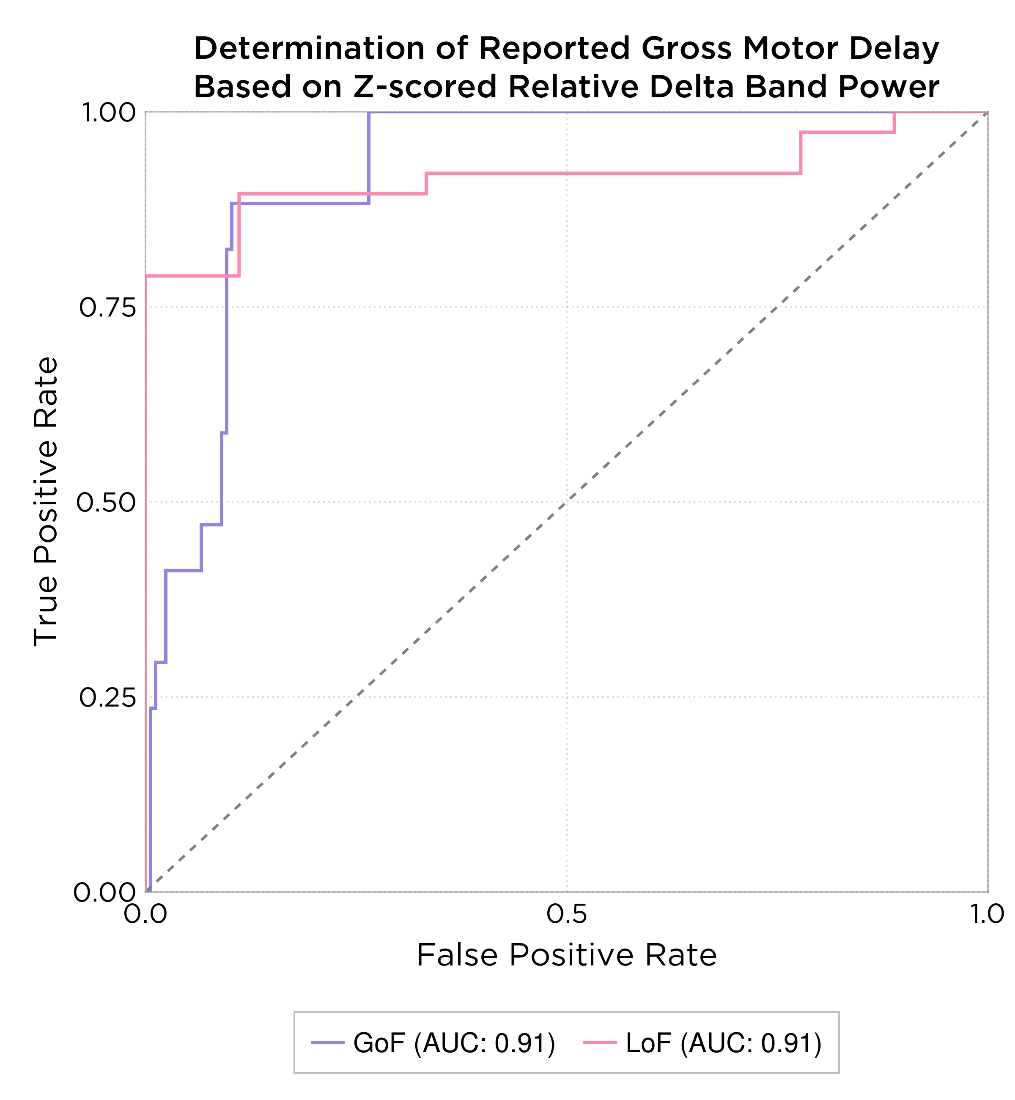

**Supplemental Figure 7.** **Model fit for model of gross motor developmental delay by z-scored relative delta band power.** ROC curve for model, split by variant classification. GoF AUC: 0.91; LoF AUC: 0.91.

**Supplemental Tables**

**Supplementary Table 1. Control recording and corresponding subject counts by age bin.**

| **Age Bin** | **Recording Count** | **Subject Count** |
| --- | --- | --- |
| **0-2mo** | 63 | 50 |
| **2-6mo** | 81 | 60 |
| **6mo-1yr** | 89 | 61 |
| **1-2yr** | 213 | 148 |
| **2-4yr** | 271 | 187 |
| **4-8yr** | 361 | 274 |
| **8-16yr** | 626 | 469 |

**Supplementary Table 2. Gross motor developmental milestone limits.**

Source abbreviations: WHO: World Health Organization^46^; FRCPC: Fellows of the Royal College of Physicians of Canada meta-analysis^47^; Adolph^48^.

| **Milestone** | **Limit (months)** | **Source** |
| --- | --- | --- |
| **Ability to sit unsupported** | 8.0 | WHO |
| **Ability to stand** | 10.1 | WHO |
| **Ability to crawl** | 11.3 | WHO |
| **Ability to walk with assistance** | 11.8 | WHO |
| **Ability to stand alone** | 14.4 | WHO |
| **Ability to walk** | 15.3 | WHO |
| **Ability to roll (back-to-front)** | 9.0 | FRCPC |
| **Ability to roll (front-to-back)** | 9.0 | FRCPC |
| **Ability to roll** | 9.0 | FRCPC |
| **Ability to run** | 24.0 | FRCPC |
| **Ability to sit** | 8.0 | WHO |
| **Ability to jump** | 24.0 | FRCPC |
| **Ability to pull to stand** | 12.0 | FRCPC |
| **Ability to belly crawl** | 11.3 | WHO |
| **Ability to throw** | 12.0 | FRCPC |
| **Ability to walk with assistance** | 12.0 | FRCPC |
| **Ability to cruise** | 13.5 | Adolph |

**Supplementary Table 3.  *SCN2A* cohort demographics by subject.** Subject demographics for all subjects included in **Table 1**, with one row per subject. Note, a missing *SCN2A* diagnosis age (occurs for one subject) indicates that the variant was not confirmed as causative by their treating physician.

| **Subject** | **Variant consensus score** | **Variant classification** | **Phenotype** | **SCN2A diagnosis age bin** | **Months from first reported seizure or epilepsy diagnosis to SCN2A diagnosis** | **Autism diagnosis (Y/N)** | **Median number of prescribed ASMs** | **Median number of prescribed SCBs** | **Recording count** | **Median recording age bin** |
| --- | --- | --- | --- | --- | --- | --- | --- | --- | --- | --- |
| **1** | GoF | GoF | EO | 0-2mo | 0.624 | N | 3.0 +/- 0.5 | 1.5 +/- 1.0 | 9 | 0-2mo |
| **2** | Possibly-GoF | GoF | EO | 0-2mo | 1.512 | N | 2.0 +/- 0.0 | 2.0 +/- 0.0 | 60 | 0-2mo |
| **3** | Likely-GoF | GoF | EO | 0-2mo | 1.8 | N | 2.0 +/- 2.0 | 1.0 +/- 0.0 | 13 | 2-6mo |
| **4** | GoF | GoF | EO | 0-2mo | 0.36 | N | 1.0 +/- 1.0 | 0.0 +/- 0.0 | 6 | 6mo-1yr |
| **5** | Likely-GoF | GoF | EO | 8-16yr | 231.924 | N | 1.0 +/- 1.0 | 1.0 +/- 0.0 | 6 | 8-16yr |
| **6** | GoF | GoF | EO | 1-2yr | 13.896 | N | 2.0 +/- 1.0 | 1.0 +/- 0.0 | 41 | 2-6mo |
| **7** | GoF | GoF | EO | 8-16yr | 129.156 | N | 1.0 +/- 2.0 | 1.0 +/- 0.5 | 3 | 8-16yr |
| **8** | Likely-GoF | GoF | EO | 8-16yr | 114.66 | Y | 2.0 +/- 2.0 | 1.0 +/- 1.0 | 16 | 4-8yr |
| **9** | GoF | GoF | EO | 6mo-1yr | 10.248 | N | 5.0 +/- 3.0 | 1.0 +/- 1.0 | 28 | 2-4yr |
| **10** | Possibly-GoF | GoF | EO | 2-6mo | 2.196 | Y | 2.0 +/- 0.5 | 1.0 +/- 1.0 | 24 | 0-2mo |
| **11** | Likely-GoF | GoF | EO | 2-4yr | 28.524 | N | 3.0 +/- 3.0 | 1.0 +/- 2.0 | 57 | 2-6mo |
| **23** | Likely-LoF | LoF | EO | 2-4yr | 40.308 | Y | 1.0 +/- 0.0 | 1.0 +/- 0.0 | 5 | 2-4yr |
| **19** | LoF | LoF | LO | 8-16yr | 45.276 | Y | 1.0 +/- 0.0 | 1.0 +/- 0.0 | 20 | 8-16yr |
| **20** | LoF | LoF | LO | 2-4yr | -12.06 | N | 1.0 +/- 0.0 | 0.0 +/- 0.0 | 21 | 4-8yr |
| **22** | Possibly-LoF | LoF | LO | N/A | N/A | Y | 3.0 +/- 2.0 | 0.0 +/- 0.0 | 43 | 4-8yr |
| **24** | Possibly-LoF | LoF | LO | 8-16yr | 147.048 | Y | 1.0 +/- 0.0 | 1.0 +/- 1.0 | 1 | 8-16yr |
| **25** | LoF | LoF | LO | 1-2yr | -9.984 | Y | 1.0 +/- 0.0 | 0.0 +/- 0.0 | 1 | 1-2yr |
| **26** | LoF | LoF | LO | 2-4yr | 2.424 | Y | 2.0 +/- 0.25 | 0.0 +/- 1.0 | 2 | 4-8yr |
| **27** | LoF | LoF | LO | 2-4yr | 1.776 | Y | 2.0 +/- 0.0 | 0.0 +/- 0.0 | 15 | 4-8yr |
| **12** | LoF | LoF | LOIS | 1-2yr | 9.78 | Y | 1.0 +/- 1.0 | 0.0 +/- 0.0 | 33 | 4-8yr |
| **13** | Likely-LoF | LoF | LOIS | 1-2yr | -8.712 | N | 3.0 +/- 1.0 | 0.0 +/- 0.0 | 16 | 2-4yr |
| **14** | Likely-LoF | LoF | LOIS | 6mo-1yr | 0.36 | Y | 3.0 +/- 1.0 | 0.0 +/- 0.0 | 13 | 4-8yr |
| **15** | Possibly-LoF | LoF | LOIS | 8-16yr | 132.012 | Y | 1.0 +/- 0.0 | 0.0 +/- 0.0 | 11 | 2-4yr |
| **16** | Possibly-LoF | LoF | LOIS | 4-8yr | 40.08 | N | 1.0 +/- 1.0 | 0.0 +/- 0.0 | 22 | 4-8yr |
| **17** | Likely-LoF | LoF | LOIS | 1-2yr | 5.016 | N | 3.5 +/- 2.0 | 0.0 +/- 0.0 | 4 | 2-4yr |
| **18** | Possibly-LoF | LoF | LOIS | 6mo-1yr | 2.304 | N | 1.0 +/- 0.0 | 0.0 +/- 0.0 | 2 | 2-4yr |
| **21** | Possibly-LoF | LoF | LOIS | 6mo-1yr | 0.72 | N | 3.0 +/- 0.0 | 0.0 +/- 0.0 | 5 | 6mo-1yr |
| **28** | Likely-LoF | LoF | LOIS | 1-2yr | 2.112 | N | 2.0 +/- 2.0 | 0.0 +/- 0.0 | 16 | 1-2yr |
